# Pattern of Thyrocardiac Disease among Patients with Hyperthyroidism in Hong Kong

**DOI:** 10.1101/2022.07.06.22277145

**Authors:** Daniel Kwun-Hung Lau, Ka-Ho Mok, Cheuk-Wing Ng, Cho-Pui Lam, Zhiwei Liu, Bai-Lin Li, Jian-Hua Lao, Shing-Fung Ma

## Abstract

This research aims to investigate the association between hyperthyroidism and age, sex, and infraction of the myocardium in the Hong Kong population in order to detect hyperthyroidism-induced infraction earlier (e.g., thyrocardiac disease) through chemiluminescent microparticle immunoassay (CMIA). A total of 7616 random blood specimens from males and females were collected and examined by Architect i immunoassay platform to obtain quantitative laboratory values of Free FT4, TSH, and hs-cTnI in samples serum differentiating into overt and subclinical hyperthyroidism cases. The age was between 19 and 81 years old. In this research, for population characteristics, 57 cases confirmed as hyperthyroidism (e.g. FT4 > 1.48 ng/dL, according to Abbott’s suggested reference interval) were used for analysis and 22 cases were examined in the laboratory. The highest number of hyperthyroidisms including overt and subclinical cases in female was 43 (75.44%), rather than males. For prevalence data of hyperthyroidism between September 2019 to January 2020 in Hong Kong, the range was from 0.62% to 0.93% (Median percentage = 0.75%). The research will explore the in-depth impact of gender difference on thyrocardiac disease, risk of thyrocardiac disease by age group, and the possible relationship between FT4 and hs-cTnI.

## 1. Introduction

For thyrocardiac disease, Mulatu (2019) stated that the existence of combination of thyroid toxicity and heart disease is found in an individual patient. The frequent and clinical signs of thyrocardiac disease are hypertension, atrial fibrillation, pulmonary hypertension and dilated cardiomyopathy (DCM), causing a damage to the heart muscles.[23] Hyperthyroidism clinically means overactivity of the thyroid gland. Besides, thyrotoxicosis clinically means raised level of blood T4 due to high metabolic rate. [28] Hyperthyroidism can also be detected by elevation of FT4 level due to the overactive thyroid gland. The increased thyroid hormone (e.g., FT4 or T4) increases the accumulation of intracellular calcium, resulting an increase in cardiac output. [24] So that, the effects of thyroid hormones will become a clinical change in cardiac rhythmicity or of heart muscle function. In thyrocardiac disease, heart disease is caused by hypertension, atrial fibrillation, pulmonary hypertension and dilated cardiomyopathy (DCM). The contraction of heart muscle is come from excitation-contraction coupling in cardiac muscle in which depends on calcium ions to play an important role for the intra-sarcomeric movement in muscular excitation-contraction coupling. [25] Globally, Tunbridge et al indicated that the average prevalence of hyperthyroidism during 1982 is around 1.0% in a population study performed in England. [26] In one of the other studies [27], Vanderpump et al found that the ratio of female to male tends to be 10:1.1. However, in a cohort study analyzed more than 20 years, the incidence of hyperthyroidism is 0.8%. Moreover, hyperthyroidism is commonly found among the younger age group as easily. [27]

Heart diseases were the third leading cause of deaths accounting for 13.2% of all deaths in Hong Kong in 2015, with about 77600 in-patient discharges and in-patient deaths in all hospitals, and 6190 deaths registered. There were 99.5 deaths due to heart disease per 100,000 males and 72.4 deaths due to heart disease per 100,000 females in 2015. [3] Women with inactivated X chromosomes often suffer from thyroid diseases because of the X chromosome encoding for many immune-related genes like FOXIP3 and CD40, which code for the CD40L receptor and are linked to thyroid disease (e.g., Graves’ disease). [4,5] In addition to thyroid hormone, sex hormone binding hormone globulin (SHBG) plays a key role in immune function. [6] Also, a study of Fisher Rat Thyroid Cell Line cells (FRTL-5) demonstrated estrogen receptors together with the ability of estradiol to reduce the NIS gene expression and increase cell growth over time, suggesting a risk of goiter development. [7]

For the prevalence of overt hyperthyroidism, this study compared Hong Kong’s results with those from other global regions. [8] In different countries, the range should be between 0.2% and 1.3% using random sampling. According to some publications [4,5], the effects of gonadal hormones like prolactin and estrogen have been suggested as a possible cause of overt hyperthyroidism. Furthermore, direct effects of estrogen on thyroid tissue in women can lead to thyroid goiter, nodule, and cancer, eventually resulting in excessive thyroid hormone (e.g., FT4) that causes hyperthyroidism by overproducing thyroid hormones. [4,5] For this study, the following hypothesis and the research questions will be tested. First of all, high FT4 and high hs-cTnI values will be obtained from the patient with hyperthyroidism. Secondly, hyperthyroidism is related and associated to female in Hong Kong. Thirdly, from the designated and categorized age groups, which of the age group will become the most dangerous and highest chance of suffering from thyrocardiac disease in Hong Kong.

## 2. Materials and methods

Based on the scientific-evidence findings on thyrocardiac disease [1,2], high death rate of heart disease in Hong Kong [3], and female who is easily associated with thyroid disorder such as Graves’ disease or goiter [4,5,6,7], this study constructs in-depth research to investigate the pattern or distribution for hyperthyroidism with low or intermediate risk of heart damage in the patient. In Figure 1., design of a retrospective study with 7616 patients evaluated at the private laboratory and analyzed at cooperative laboratory during September, 2019 to January, 2020 for clinical study of Hong Kong population. The patient’s serum is separated within 24 hours after blood collection, and stored at temperatures which is colder than −25°C. All study data were anonymized before analysis. The results of former study did not affect the clinical management of the referring doctors for the patients. No conflict of interest and no commercial value.

**Figure 1.**
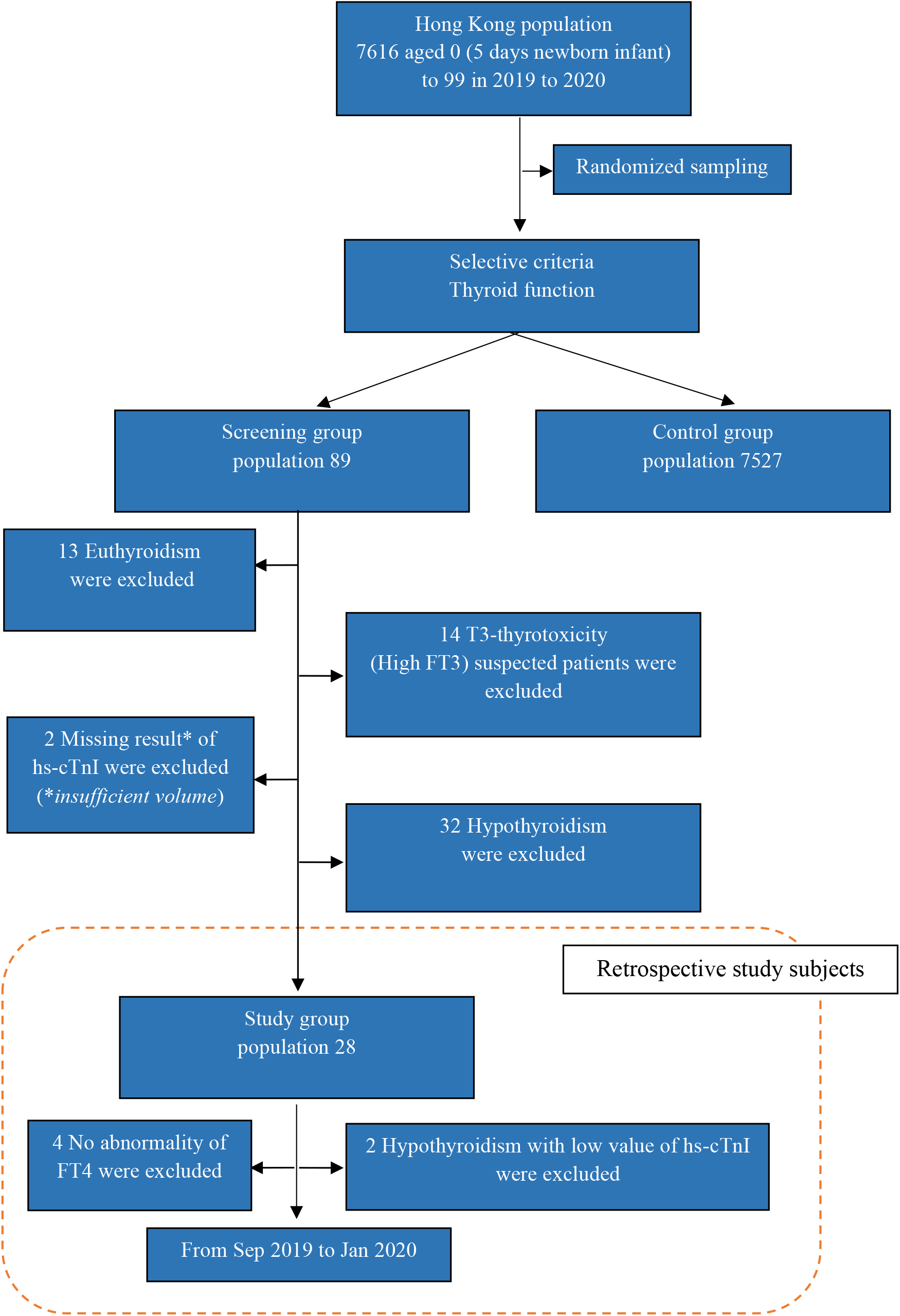
Sampling Model from randomized sampling pool to study group.

By using Chemiluminescent microparticle immunoassay (CMIA) of Architect i1000SR immunoassay analyzer to obtain quantitative laboratory values of FT4 and hs-cTnI in samples serum differentiating into overt hyperthyroidism and myocardial infraction cases. Cardiac troponin especially troponin-I is the ideal biomarker for the detection of injury from myocardial infraction. High sensitivity assay (hs-cTnI) provides an improved sensitivity and superior tissue-specificity by a comparison with other available cardiac biomarkers such as myoglobin, creatine kinase (CK), lactate dehydrogenase (LDH), and the other non-specific cardiac marker such as Aspartate Aminotransferase (AST). Two assays of reagents (Free T4; Reference code: 7K65 and STAT Troponin-I; Reference code: 2K41) are used for this research study. They are two-step immunoassay to determine the presence of FT4 and hs-cTnI in human serum using Chemiluminescent microparticle immunoassay (CMIA) methodology. [29,30]

In the beginning, the antibody-coated paramagnetic microparticles are available for the desired analytes for binding. After incubation and washing, the labeled conjugate is added to form an antibody-coated paramagnetic microparticles bound complex. This is second step. And for further incubation and washing, pre-trigger (hydrogen peroxide, H_2_O_2_) and trigger solutions (sodium hydroxide, NaOH) have to be used and added to the above reaction mixture. Finally, the resulting chemiluminescent reaction is measured as the unit of relative light units (RLUs). The RLU will be detected by Architect i1000SR optical system. The concentration of analytes will be determined by correlating with standard curve established by calibrators.

For data processing and analysis, SPSS statistical software was used to summarize the data into the format of figures and graphs for data presentation. The categorical variables were compared using Pearson chi-square test and statistically significant association was considered when p-value was < 0.05. And, regression analysis will be used to evaluate designated model of analysis. R2 is close to 1.0 to indicate statistically high association of relationship between two variables.

## 3. Results

In Figure 1., 57 cases (0.75%) were detected as hyperthyroidism (57 out of 7616 specimens) which were collected randomly from the clinic. Besides, totally 22 females and 6 males out of 57 cases successfully proceed the FT4 and hs-cTnI tests performed in the research laboratory. Totally 28 are classified into study group. However, 19 females (Mean = 2.59 ng/dL) and 3 males (Mean = 2.48 ng/dL) were matched to the criteria of hyperthyroidism (e.g. FT4 >1.48 ng/dL) and selected to be final study group. In Figure 2a. & 2b., the data is showed that the age group of 19-39 has the significantly highest number (n = 15/28; 53.6%) for suffering from hyperthyroidism and low risk of cardiovascular disease in Hong Kong population. After the age group of 19-39, the number of hyperthyroidisms has a decreasing trend between 40-49 and 60-69 age groups.

**Figure 2.**
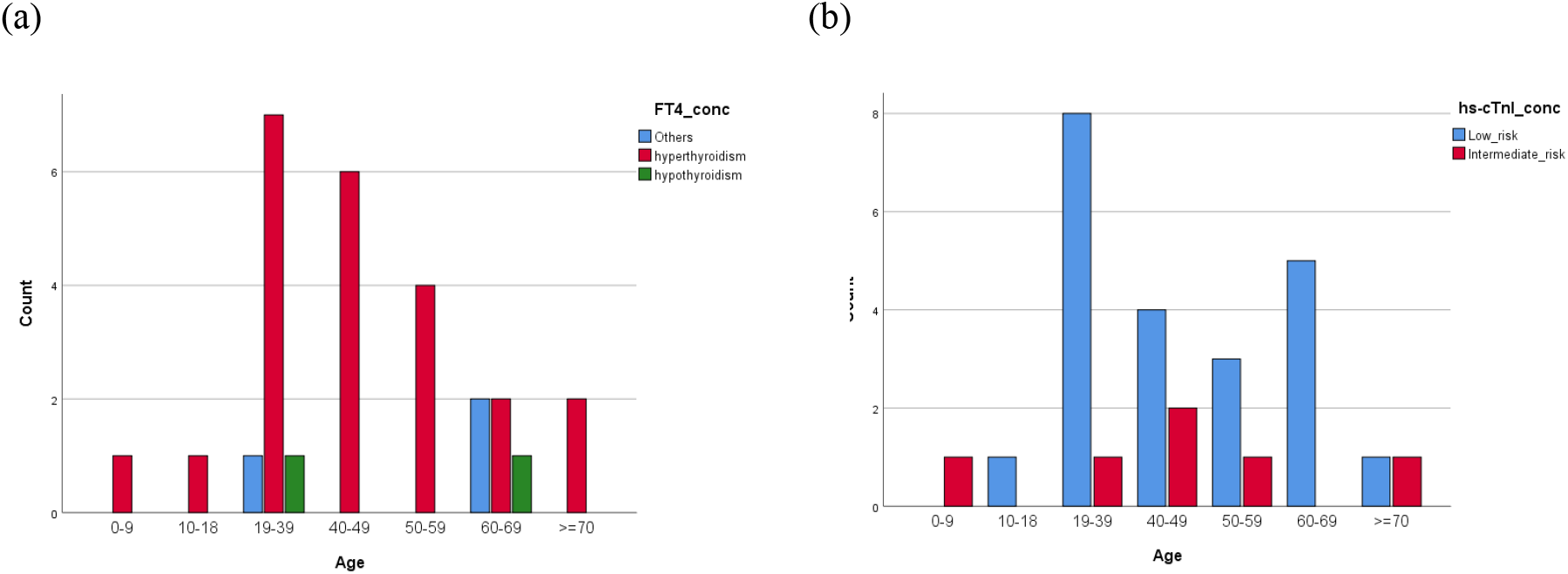
**(a)** Total 28 cases were evaluated with FT4 to separate to hyperthyroidism, hypothyroidism and others. By Chi-Square test for age and FT4, the statistical significance is resulted (*p = 0.659*). **(b)** Total 28 cases were evaluated with hs-cTnI to separate to low risk and intermediate risk for cardiovascular disease (CVD). By Pearson’s chi square test for age and hs-cTnI, the statistical significance is resulted (p = 0.287).

In Figure 3a. & 3b., the interpolation lines are obtained to observe the trend between FT4 values and ascending age groups. In the female, the highest values of FT4 and hs-cTnI in age group 40-49 from the trend line is obtained. And, for the upward trending seen from the age group 60-69 to ≥70. In Figure 4a. & 4b., the histogram and boxplot presented a clear difference among gender and age group. Female has the significantly highest number of hyperthyroidism than males. Larger sampling size if age below 50 years old becomes higher risk of getting hyperthyroidism than if age above or equal to 50 years. In Figure 4c., the boxplot showed that most of the cases (Mean = 4.07 pg/mL) are below the cutoff for intermediate risk of CVD in the final study group (n = 22). For the results of hs-cTnI test, none of both genders are higher than the reference interval of hs-cTnI ≥ 120 pg/mL.

**Figure 3.**
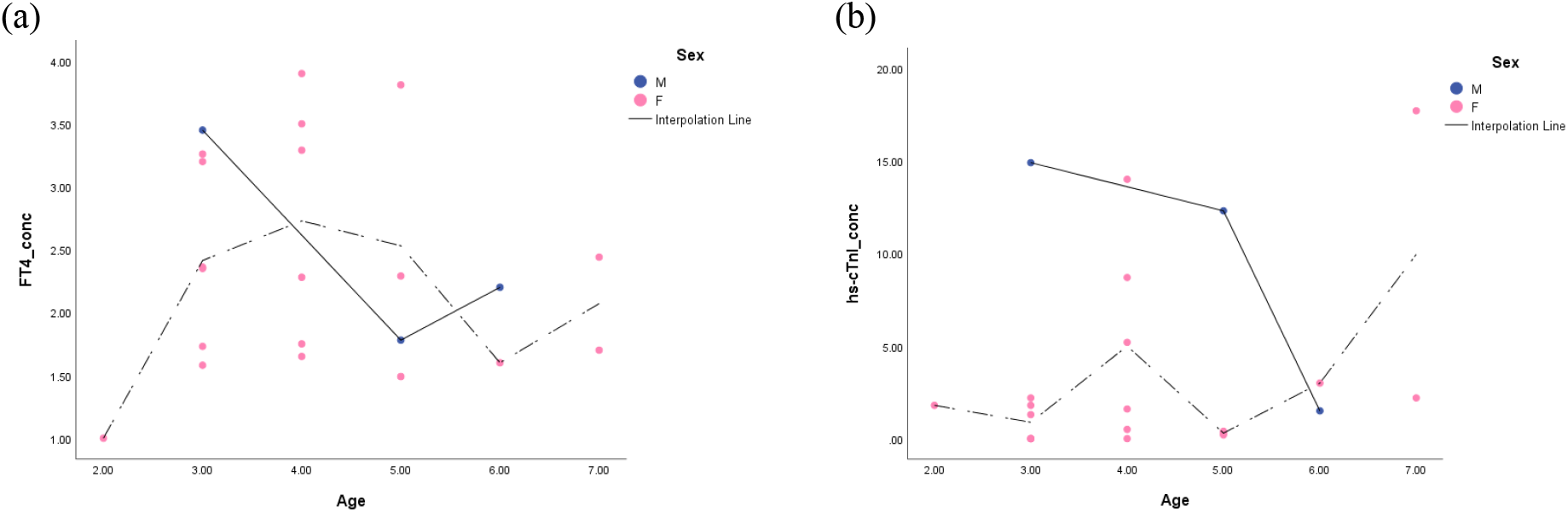
**(a)** Relationship between FT4 and age with labelling cases by sex among hyperthyroidism cases. (**b**) Relationship between hs-cTnI and age with labelling cases by sex among hyperthyroidism cases. (n = 22)

**Figure 4.**
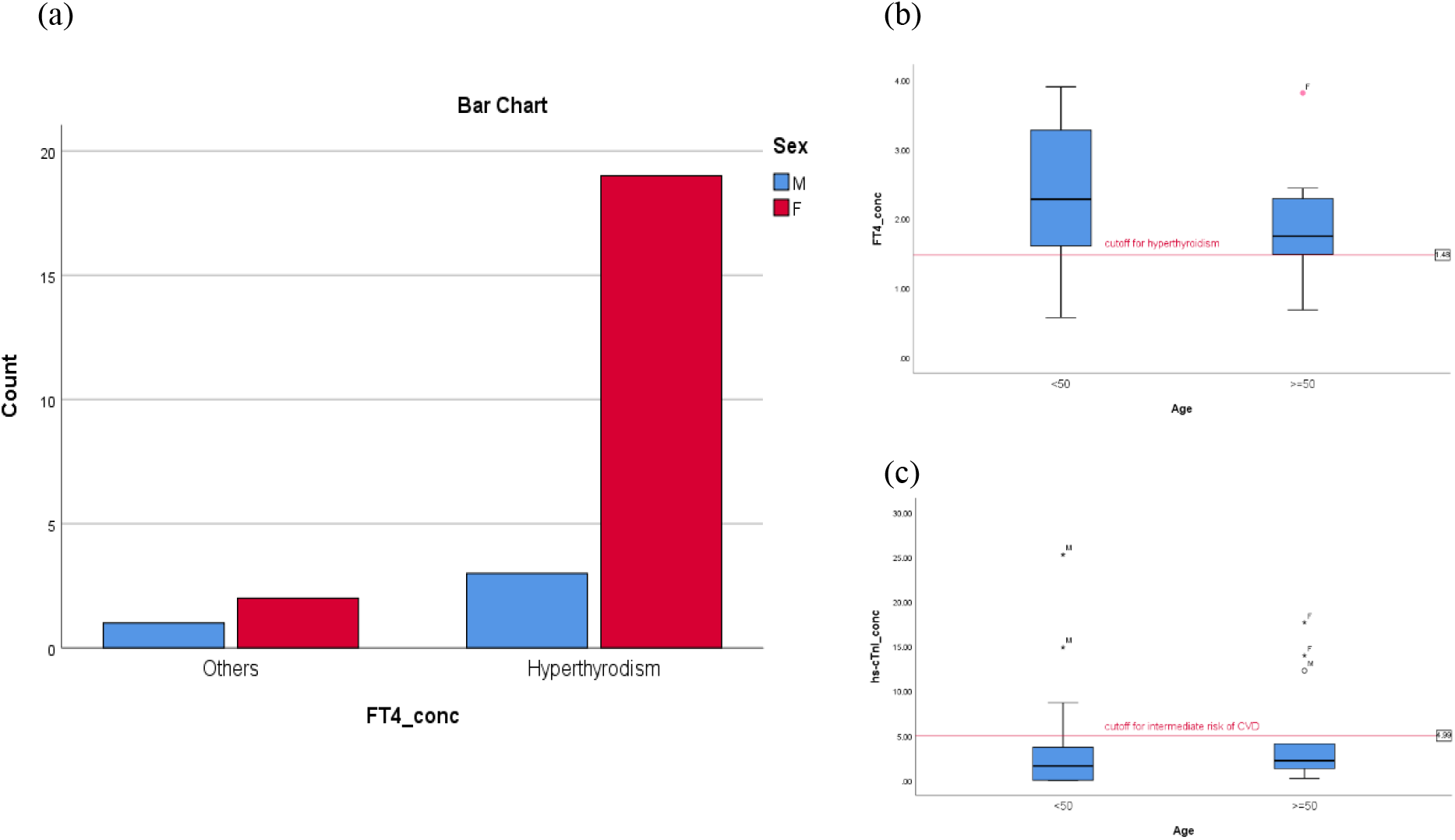
**(a)** Higher number of females suffering from hyperthyroidism than males. Hyperthyroidism: if FT4 > 1.48 ng/L; Others: normal thyroid function without any elevation of FT4 which is out of the range (0.7 – 1.48 ng/L). By using Pearson chi square test, the statistical significance value is *p = 0.383*. **(b)** Distribution for the age group (e.g. <50 and ≧50 years) against FT4 concentration. (n = 22) **(c)** Distribution for the age group (e.g. <50 and ≧50 years) against hs-cTnI concentration. (n = 22)

In Figure 5., by plotting a scatter plot, the interpolation line is obtained by 6 cases of hyperthyroidism with intermediate risk of CVD. The trend is clearly and obviously observed for continuously decreasing concentration at age group 50-59 and continuously increasing concentration at age group ≥70. The interpolation line shows the trend of the concentrations of test results under different age groups. In Figure 6a., FT4 correlation study is plotted with hs-cTnI concentration. This scatter plot shows two groups consisting of patient with hyperthyroidism & low risk of getting CVD, and hyperthyroidism & intermediate risk of getting cardiovascular disease. In Table 1., this is the data showing the mean, range, standard deviation, and variance for the resulted laboratory values of Free T4 (thyroid) and hs-cTnI (cardiac) for the different age groups by using statistical software, SPSS.

**Figure 5.**
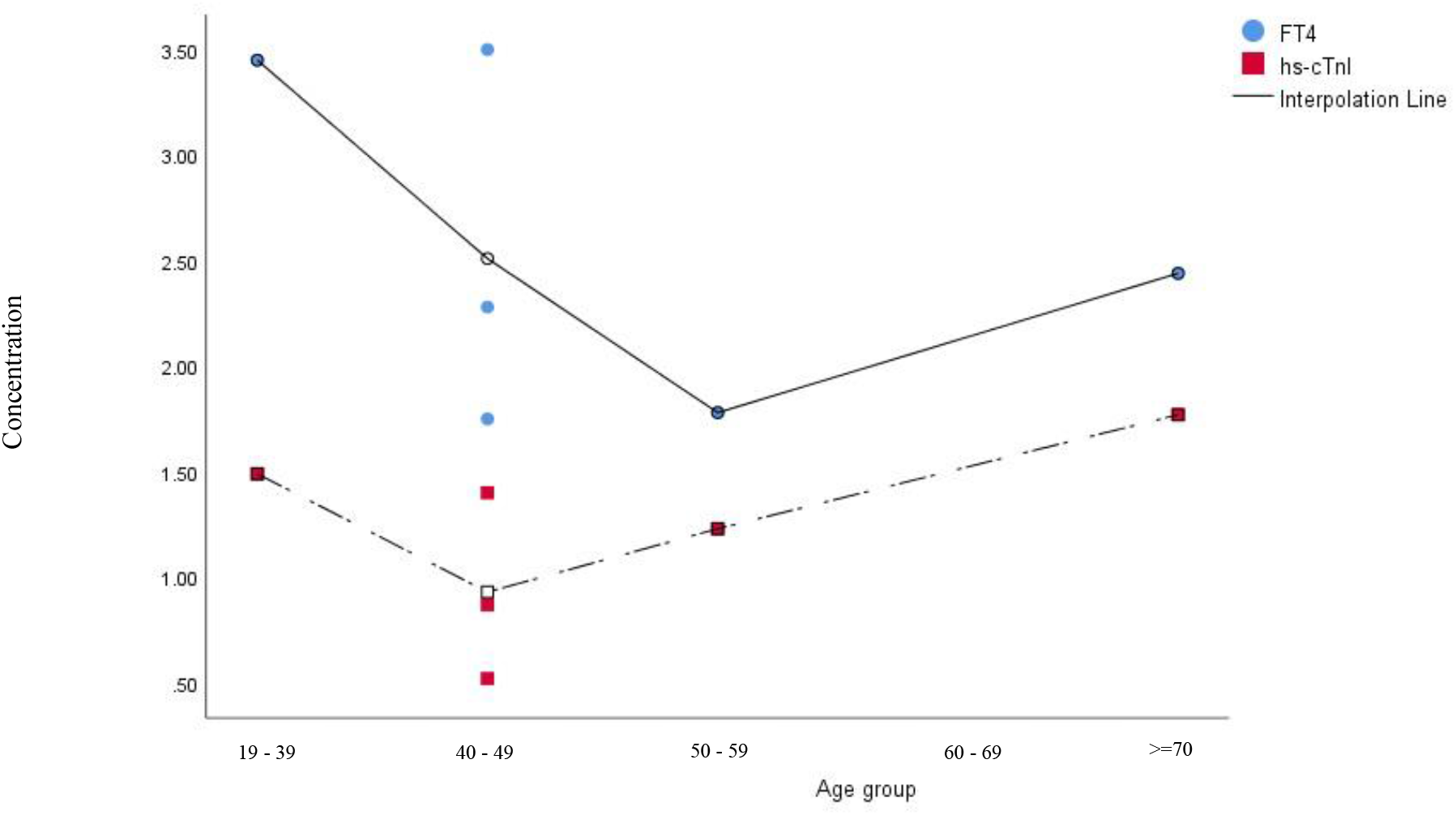
With implementation interpolation lines, the scatter plot showed the trend between the age group among hyperthyroidism and intermediate risk of cardiovascular disease concentration. (n = 6)

**Figure 6.**
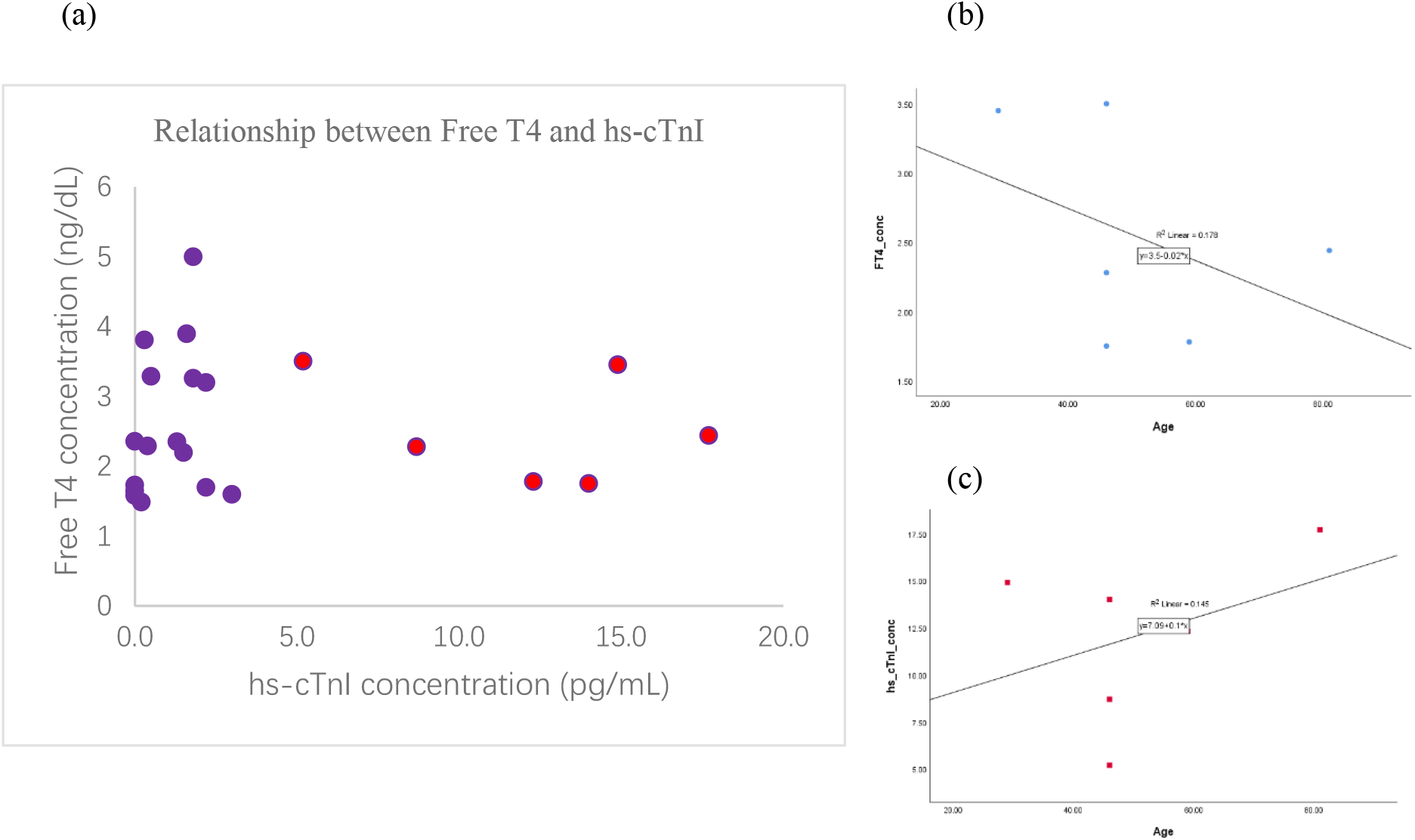
**(a)** Free T4 correlation study. Purple spot: patient with hyperthyroidism (FT4: > 1.48 ng/dL) and low risk (hs-cTnI: 0 – 4.99 pg/mL) of getting cardiovascular disease; Red spot: patient with hyperthyroidism and intermediate risk (hs-cTnI: 5 – 119 pg/mL) of getting cardiovascular disease. Correlation coefficient = -0.023. (n = 28) **(b)** FT4 against different ages. By adding a fit line and using regression analysis, R^2^ = 0.178. (n = 6). **(c)** hs-cTnI against different ages. By adding a fit line and using regression analysis, R^2^ = 0.145. (n = 6)

**Table 1.** Data showing the mean, range, standard deviation, and variance for the resulted laboratory values of Free T4 (thyroid) and hs-cTnI (cardiac) for the different age groups by using statistical software, SPSS. FT4: Free thyroxine; hs-cTnI: high sensitivity-cardiac troponin-I.

| Descriptive Statistics |  |  |  |  |  |  |  |
| --- | --- | --- | --- | --- | --- | --- | --- |
| Concentration | N | Range | Minimum | Maximum | Mean | Std. Deviation | Variance |
| FT4 | 28 | 3.32 | 0.58 | 3.90 | 2.0993 | 0.95964 | 0.921 |
| hs-cTnI | 28 | 25.30 | 0.00 | 25.30 | 4.4357 | 6.48966 | 42.116 |
| Valid N (listwise) | 25 |  |  |  |  |  |  |

## 4. Discussion

This is the first hs-cTnI based study conducted among hyperthyroidism patients in Hong Kong for evaluation of hyperthyroidism induced-myocardial infraction (this is also called thyrocardiac disease). The majority use of hs-cTnI is to detect heart damage from elevation of FT4 level circulating in blood.

### 4.1 Relationship between FT4 and hs-cTnI

The established regression models (Figure 6b. & 6c.) show that how the effect of age (which is assumed as fixed with no error) causes FT4 and hs-cTnI concentrations to change in these regression models.

Based on the slope of -0.02 (shown in Figure 6b.), the prediction and assumption are that each 1 year increases in age decreases hyperthyroidism by lowering 0.02 ng/dL of FT4 values. In contrast, based on the slope of 0.1 (shown in Figure 6c.), the prediction and assumption is that each 1 year increases in age increases myocardial infraction by increasing 0.1 pg/mL of hs-cTnI values. This might be explained by high prevalence of high FT4 values in younger age group (e.g. age below 50 years, especially in age 19-39) and high prevalence of elevated hs-cTnI values in elderly age group (e.g. age ≥70 years).

Besides, in the correlation study with two numeric variables of FT4 and hs-cTnI, there is a quick and simple summary of the direction and relationship between FT4 and hs-cTnI concentration. In the small sampling size of final study group (n=22), the correlation coefficient of -0.023 indicates a perfect negative correlation. As variable of hs-cTnI concentration increases, variable of FT4 decreases and vice versa. This might be explained elevated FT4 trigger long-term hyperthyroidism in the patient who do not have high values of hs-cTnI because acute phase of myocardial infraction is over.

In the case of patient, excessive thyroid hormones directly stimulate myocardial cells to cause increased cardiac workload. Thyroid hormone can enhance the overall effect of catecholamines, by increasing the number, affinity, cAMP activity and intracellular cAMP metabolism of adrenergic receptors on the surface of myocardial cell membrane. Furthermore, an increasing the number and excitability of cardiovascular adrenergic receptors stimulates the production of catecholamines, which are indirectly to stimulate the increase of cardiac workload and the long-term capacity. As a result, the heart is totally overloaded.

### 4.2 Impact of gender for thyrocardiac disease

Consistent with the study from other global studies [11], by using random sampling method in different countries, the result of prevalence rate for overt hyperthyroidism is between 0.2% to 1.3%. Therefore, in this research, the data for prevalence of hyperthyroidism is matched to the global study [11].

Regarding to the impact of gender on thyrocardiac disease, it is found that females are easier to suffer from thyrocardiac disease than male. For the explanation, the effects of female gonadal hormones such as estrogen and prolactin, which triggers exceptional secretion of biologically active thyroid hormone such as FT4 to cause hyperthyroidism, will finally develop thyroid goiter, multinodular goiter and thyroid cancer in a worst case. [4,5]

Besides, for the female, the X chromosome inactivation on thyroid gland contributes significantly to the development of autoimmune thyroid diseases. This is because the X chromosome contains a variety of immunological related genes such as CD40 and FOXIP3, which serve as genetic marker to show clinical and strong association with autoimmune thyroid diseases like Graves’ disease. [4,5,31]

### 4.3 Risk of thyrocardiac disease among age group

In terms of age group, this study estimated that young people between the age of 19 and 39 have under higher pressure. They face many problems than the other age group. For example, academic, work, family and housing are all the sources of stress for them. [13,14] Under stress, the body releases cortisol called stress hormone. Then, excessive cortisol causes an interference to thyroid hormone production. This stimulates the thyroid gland to work harder to produce excessive thyroid hormone in blood circulation. [15] Findings with other studies, for the elderly, most of them are retired by comparing with 19-39 age group. Hence, they may have lower level of cortisol leading to less stimulation of thyroid. [17] However, the body condition is getting worst progressively when aging. Problems with arrhythmia are much more common in older adults than younger people. And for adults aged 65 and older, they are more likely to suffer from cardiovascular disease than younger people because of the naturally metabolic changes in the heart and blood vessels. [18]

### 4.4 Limitation

For the elevated FT4 values in patient with hyperthyroidism were not closely associated with the elevated hs-cTnI values to interpret the progression of thyrocardiac disease. This might be due to relatively small sample size of diagnosed patients in this research study and needs further study by using a larger scale of population. This might not represent the true picture in the community.

## 5. Conclusion

Hyperthyroidism is proved as a higher risk to develop myocardial infraction (MI) causing thyrocardiac disease, which is similar to the other case studies. [21,22] In this research study, two factors are investigated including gender in female and age group of 19-39 for higher risk of suffering from thyrocardiac disease to develop MI. In terms of gender, female is more likely to suffer from hyperthyroidism than male, having higher chance to develop MI. Finally, the age is not the major factor affecting incidence rate of thyrocardiac disease but the young to middle age group population such as aged from 19 to 39, having a high risk to develop thyrocardiac disease.

In this research, the study will be further investigating overt and subclinical hyperthyroidism separately by different kinds of blood cardiac markers. It is assumed that overt hyperthyroidism would be more severe since the level of FT4 in overt hyperthyroidism, which is last-longer, is higher than that in subclinical hyperthyroidism, leading to severe cardiac damage. Besides, the analyzed data indicated that the females aged 19-39 to have an increased risk of hyperthyroidism, possibly causing development of MI. The data provided a vital and statistical foundation for considering and studying early detection and management for hyperthyroidism to reduce and minimize the risk for thyrocardiac disease.

## Supporting information

Supplemental table 1_modified

## Data Availability

All data produced in the present work are contained in the manuscript

## 7. Ethical consideration

This study was performed following the Declaration of Helsinki and Good Clinical Practice principles. The study protocol was reviewed and approved by the Institutional Review Board of the private laboratory (IRB number HKPHC202111-001).

## 9. Acknowledgements

The authors thank Dr. Yvonne Kam N.W. at Department Clinical Oncology, The University of Hong Kong (HKU), Dr. Daniel Tam C.C. at Department of Life and Health Sciences, HKU School of Professional and Continuing Education (HKU SPACE) and Mr. Jeffrey Lau H.Y. MBBS at Department of Medicine, HKU for their advice regarding this manuscript improvement. Technical support was provided by Biomedical Laboratory at Department of Health and Life Sciences, Hong Kong Institute of Vocational Education (IVE, Sha Tin). No fund support and all researchers have no conflict of interests in this research.

